# Mapping psychological distress, depression and anxiety measures to adolescent AQoL-6D utility using data from a sample of young people presenting to primary mental health services

**DOI:** 10.1101/2021.07.07.21260129

**Authors:** Matthew P Hamilton, Caroline Gao, Kate M Filia, Jana M Menssink, Sonia Sharmin, Nic Telford, Helen Herrman, Ian B Hickie, Cathrine Mihalopoulos, Debra J Rickwood, Patrick D McGorry, Sue M Cotton

## Abstract

**Background:** Health utility data are rarely routinely collected in mental helath services. Mapping models that predict health utility from other outcome measures are typically derived from cross-sectional data but often used to predict longitudinal change.

**Objective:** We aimed to develop models to map six psychological measures to adolescent Assessment of Quality of Life – Six Dimensions (AQOL-6D) health utility for youth mental health service clients and assess the ability of mapping models to predict longitudinal change.

**Methods:** We recruited 1107 young people attending Australian primary mental health services, collecting data at two time points, three months apart. Five linear and three generalised linear models were explored to identify the best mapping model. Ten-fold cross-validation using ***R*^2^**, root mean square error (RMSE) and mean absolute error (MAE) were used to compare models and assess predictive ability of six candidate measures of psychological distress, depression and anxiety. Linear / generalised linear mixed effect models were used to construct longitudinal predictive models for AQoL-6D change.

**Results:** A depression measure (Patient Health Questionnaire-9) was the strongest independent predictor of health utility. Linear regression models with complementary log-log transformation of utility score were the best performing models. Between-person associations were slightly larger than within-person associations for most of the predictors.

**Conclusions:** Adolescent AQoL-6D utility can be derived from a range of psychological distress, depression and anxiety measures. Mapping models estimated from cross-sectional data can approximate longitudinal change but may slightly bias health utility predictions.

**Data:** Replication code and model catalogues are available at: https://doi.org/10.7910/DVN/DKDIB0.

## 1 Introduction

Quality adjusted life years (QALYs) are indices of outcome that inform public health policy in many countries [1]. One of the main uses of QALYs is as the measure of benefit in Cost-Utility Analyses (CUAs). Compared to economic evaluations that use other outcome measures, CUAs facilitate comparisons of the value for money claims of interventions for different health conditions. CUAs have this advantage because of the generic nature of health outcomes measured by QALYs and the existence of well understood policymaker willingness to pay thresholds for QALYs.

The “quality” in QALYs is measured via the use of multi-attribute utility instruments (MAUIs), where domains of quality of life measured by a questionnaire are weighted using people’s preferences [2]. This approach produces a single health utility value for each individual at each measured health state, anchored on a scale where 0 represents a state equivalent to death and 1 represents perfect health. Health utilities can be converted to QALYs by weighting the duration (the “years” part of QALYs) each individual spends in each health state.

MAUIs are regularly used in research studies such as clinical trials and epidemiological surveys, but rarely feature in routine data collection by mental health services. In the absence of direct measurement, mapping analysis has been developed to predict health utility from standard health status measurements [3, 4]. MAUIs such as the Assessment of Quality of Life – Eight Dimensions (AQoL-8D [5]) have been shown to be sensitive to psychological measures [6] and can be mapped to [7] using measures of psychological distress (measured using Kessler Psychological Distress Scale – 10 items, K10 [8]) and depression and anxiety symptoms (measured using Depression, Anxiety, and Stress Scale – 21 items, DASS-21 [9]). Measures of anxiety (Generalised Anxiety Disorder Scale (GAD-7; [10]) and depression (Patient Health Questionnaire-9 (PHQ-9; [11]) have recently been used to map to other MAUIs [12]. However, it is unclear which mental health measures are the most predictive of health utility and existing algorithms developed for adult [7] or child [13] general populations are of questionable appropriateness for predicting health utility in clinical youth mental health samples.

Currently available mapping algorithms are largely derived from cross-sectional data and assume that the associations between psychological measurements and health utility are time-invariant. Therefore, the between-person association (variations in mental health symptoms associated with variations in health utility observed cross-sectionally in a population) can be applied to estimate the within-person association (changes in mental health symptoms associated with changes in health utility over time). However, this time invariant assumption may not be true (for example, health utility measures may be less sensitive to change compared with measures of mental health symptoms).

We are developing a computational model of the systems shaping the mental health of young people [14]. To enable that model to be used in cost-utility analyses of primary youth mental health services, we aimed to use data from a sample of help-seeking young people to: (i) identify the best mapping regression models to predict adolescent weighted AQoL-6D utility from six candidate measures of psychological distress, depression and anxiety; and (ii) assess ability of the mapping algorithms to predict longitudinal (three-month) change.

## 2 Methods

### 2.1 Ethics approval

The study was approved by the University of Melbourne’s Human Research Ethics Committee and the local Human Ethics and Advisory Group (1645367.1)

### 2.2 Sample and setting

This study forms part of a youth outcomes measurement research program, and the study sample has previously been described [15]. Briefly, young people aged 12 to 25 years who presented for a first appointment for mental health or substance use related issues were recruited from three metropolitan and two regional Australian youth-focused primary mental health clinics (headspace centres) between September 2016 to April 2018. Sample characteristics are similar to previous descriptions of headspace clients, with slight differences in age (less aged 12-14, more aged 18-20), cultural background (more Culturally and Linguistically Diverse and less Aboriginal and Torres Strait Islander young people), sexuality (fewer heterosexual clients) and housing (more in unstable accommodation) [15].

### 2.3 Measures

We collected data on health utility, six candidate predictors of health utility including psychological distress, depression and anxiety measures as well as demographic, clinical and functional population information.

#### 2.3.1 Health utility

We assessed health utility using the adolescent version of the Assessment of Quality of Life – Six Dimension scale (AQoL-6D; [16]) MAUI. It was selected due to its validity for use in adolescents, the relevance of its domains for a clinical mental health sample [6] and its acceptable participant time-burden. The AQoL-6D instrument contains 20 items across the six dimensions of independent living, social and family relationship, mental health, coping, pain and sense. Health utility scores were calculated using a published algorithm (available at https://www.aqol.com.au/index. php/aqolinstruments?id=92), using Australian population preference weights. The distribution of AQoL-6D instrument item specific scores within our sample has been described in a previous study [17].

#### 2.3.2 Candidate predictors

Data from six measures of psychological distress (one measure), depression (two measures) and anxiety (three measures) symptoms were used as candidate predictors to construct mapping models. These measures are widely used in clinical mental health services or clinically relevant to the profiles of young people seeking mental health care. The Kessler Psychological Distress Scale (K6; [8]) was used to measure psychological distress over the last 30 days. It includes six items (nervousness, hopelessness, restlessness, sadness, effort, and worthlessness) of the 10 item version of this measure, K10. We used the US scoring system, in which each item uses a scale that spans from 0 (“none of the time”) to 4 (“all of the time”).

The Patient Health Questionnaire-9 (PHQ-9; [11]) and Behavioural Activation for Depression Scale (BADS; [18]) were used to measure degree of depressive symptomatology. PHQ-9 includes nine questions measuring the frequency of depressive thoughts (including self-harm/suicidal thoughts) as well as associated somatic symptoms (e.g., sleep disturbance, fatigue, anhedonia, appetite, psychomotor changes) in the past two weeks. PHQ-9 uses a four-point frequency scale ranging from 0 (“Not at all”) to 3 (“Nearly every day”). For the PHQ-9 a total score is derived (0-27) with higher scores depicting greater symptom severity. BADS measures a range of behaviours (activation, avoidance/rumination, work/school impairment as well as social impairment) reflecting severity of depression. BADS includes 25 questions on behaviours over the past week, scored on a seven-point scale ranging from 0 (“Not at all”) to 6 (“Completely”). A total score is derived for the BADS (0-150) as well as subscale scores, with higher scores indicating greater activation.

The Generalised Anxiety Disorder Scale (GAD-7; [10]), Screen for Child Anxiety Related Disorders (SCARED; [19]) and Overall Anxiety Severity and Impairment Scale (OASIS; [20]) were used to measure anxiety symptoms. GAD-7 measures symptoms such as nervousness, worrying and restlessness, over the past two weeks using seven questions, with a four-point frequency scale ranging from 0 (“Not at all”) to 3 (Nearly every day”). A total score is calculated with scores ranging from 0 to 21 and higher scores indicating more severe symptomatology. SCARED is an anxiety screening tool designed for children and adolescents which can be mapped directly on specific Diagnostic and Statistical Manual of Mental Disorders (DSM-IV-TR) anxiety disorders including generalised anxiety disorder, panic disorder, separation anxiety disorder and social phobia. It includes 41 questions on a three-point scale of 0 (“Not true or hardly ever true”), 1 (“Somewhat True or Sometimes True”) and 2 (“Very true or often true”) to measure symptoms over the last three months. A total score is derived with scores ranging from 0-82, with higher scores indicative of the presence of an anxiety disorder. The OASIS was developed as a brief questionnaire to measure severity of anxiety and impairment in clinical populations. The OASIS includes five questions about frequency and intensity of anxiety as well as related impairments such as avoidance, restricted activities and problems with social functioning over the past week. Total scores range from 0-20 with higher scores depicting more severe symptomatology.

#### 2.3.3 Population characteristics

We collected self-reported demographic measures (age, gender, sex at birth, education and employment status, languages spoken at home and country of birth). We also collected clinician or research interviewer assessed measures of mental health including primary diagnosis, clinical stage [21] and functioning (measured by the Social and Occupational Functioning Assessment Scale (SOFAS) [22]).

### 2.4 Procedures

Eligible participants were recruited by trained research assistants and responded to the questionnaire via a tablet device. Participants’ clinical characteristics were obtained from clinical records and research interview. At three-months post-baseline, participants were contacted in person or by telephone, to complete a follow-up assessment.

### 2.5 Statistical analysis

We undertook a three-step process to: i) gain insight into our dataset and measures; ii) specify models and assess model performance using cross-sectional data; and iii) specify longitudinal models and assess whether modelling assumptions were met.

#### 2.5.1 Pre-modelling steps

Basic descriptive statistics characterised the cohort in terms of baseline demographics and clinical variables. Pearson’s Product Moment Correlations (*r*) were used to determine the relationships between candidate predictors and AQoL-6D utility score.

#### 2.5.2 Cross-sectional model specification and assessment

Good practice guidance on mapping studies [4] does not advocate specific model types as the appropriate choice will vary depending on factors that include the health utility measure being mapped to, the applicable health condition and target population, the clinical variables used as predictors and the intended use of the mapping algorithm. Compared to the EQ-5D utility measures most commonly used in the mapping literature [23], AQOL-6D has better dimensional overlap with mental health measures [6] and is derived from a greater number of questionnaire items (which can generate more continuous utility score distributions). The types of models we explored reflected these considerations.

We used a cross-section of our dataset (baseline measures only) to explore appropriate type(s) of models to use, compare the relative predictive performance of candidate predictors, and identify other potential risk factors associated with quality of life independent of mental health measures.

As adolescent AQoL-6D utility score is normally left skewed and constrained between 0 and 1, ordinary least squares (OLS) models with different types of outcome transformations (such as log and logit) have been previously used in mapping [3]. Similarly, generalised linear models (GLMs) address this issue via modelling the distribution of the outcome variable and applying a link function between the outcome and linear combination of predictors [24]. Beta regressions, which can be considered a special form of GLMs, are another popular strategy for modelling health utility [25]. We chose to explore OLS, GLM and beta models with commonly adopted transformation algorithms. The models we selected for comparison were OLS regression with log, logit, log-log (*f* (*y*) = *−log*(*−log*(*y*))) and clog-log (*f* (*y*) = *log*(*−log*(1 *− y*))) transformation; GLM using Gaussian distribution with log link; and beta regression with logit and clog-log link. For each candidate model type, we evaluated the modelling performance and predictive ability of a univariate model using the candidate predictor with the highest Pearson correlation coefficient with utility scores. Ten-fold cross-validation was used to compare model fitting using training datasets and predictive ability using testing datasets using three indicators including R^2^, root mean square error (RMSE) and mean absolute error (MAE) [26, 27]. After identifying the best performing model type, we used 10-fold cross-validation to compare predictive ability of six mental health measures in predicting AQol-6D (one model for each candidate predictor).

To evaluate whether candidate predictors could independently predict utility scores, we added a range of independent variables to each of the six models that included participants’ age, sex at birth, clinical stage, cultural and linguistic diversity, education and employment status, primary diagnosis, region of residence (whether metropolitan – based on location of attending service) and sexual orientation. Functioning (as measured by SOFAS) was also included in each model to evaluate whether it can jointly predict utility with clinical symptom measurements.

#### 2.5.3 Longitudinal modelling and assumption testing

After identifying the best mapping regression model(s) for predicting between person change, we established longitudinal models to predict within person change. This was achieved using generalised linear mixed-effect models (GLMMs) including both the baseline and follow-up data. All records with complete baseline data (with or without follow-up data) were included.

Bayesian linear mixed models were used to avoid common convergence problems in frequentist tools [28]. Linear mixed effect models (LMMs) can be fitted in the same framework with Gaussian distribution and identify link function. Clustering at individual level is controlled via including random intercepts. Model fitting was evaluated using Bayesian R^2^ [29].

We compared the model coefficients for each predictor’s score at baseline and score change from baseline to assess the potential bias of estimating score changes within individuals using score difference between individuals.

### 2.6 Replicability and reuse

We undertook all our analyses using ***R*** 4.0.5 [30], using a wide range of packages (see Online Supplement, A.5). To aid study replication and appropriate out of sample re-use of our mapping models, we developed novel software that we have previously described [14] and distributed study code, data and documentation in online repositories (see Availability of data and materials).

## 3 Results

### 3.1 Cohort characteristics

Participants characteristics are summarised in Table 1. This study included 1068 (out of the 1073) participants with complete AQoL-6D data at baseline. This cohort predominantly comprised individuals with anxiety/depression (76.650%) at early (prior to first episode of a serious mental disorder) clinical stages (91.707%). Participant ages ranged between 12-25 with a mean age of 18.129 (SD = 3.263).

**Table 1.**
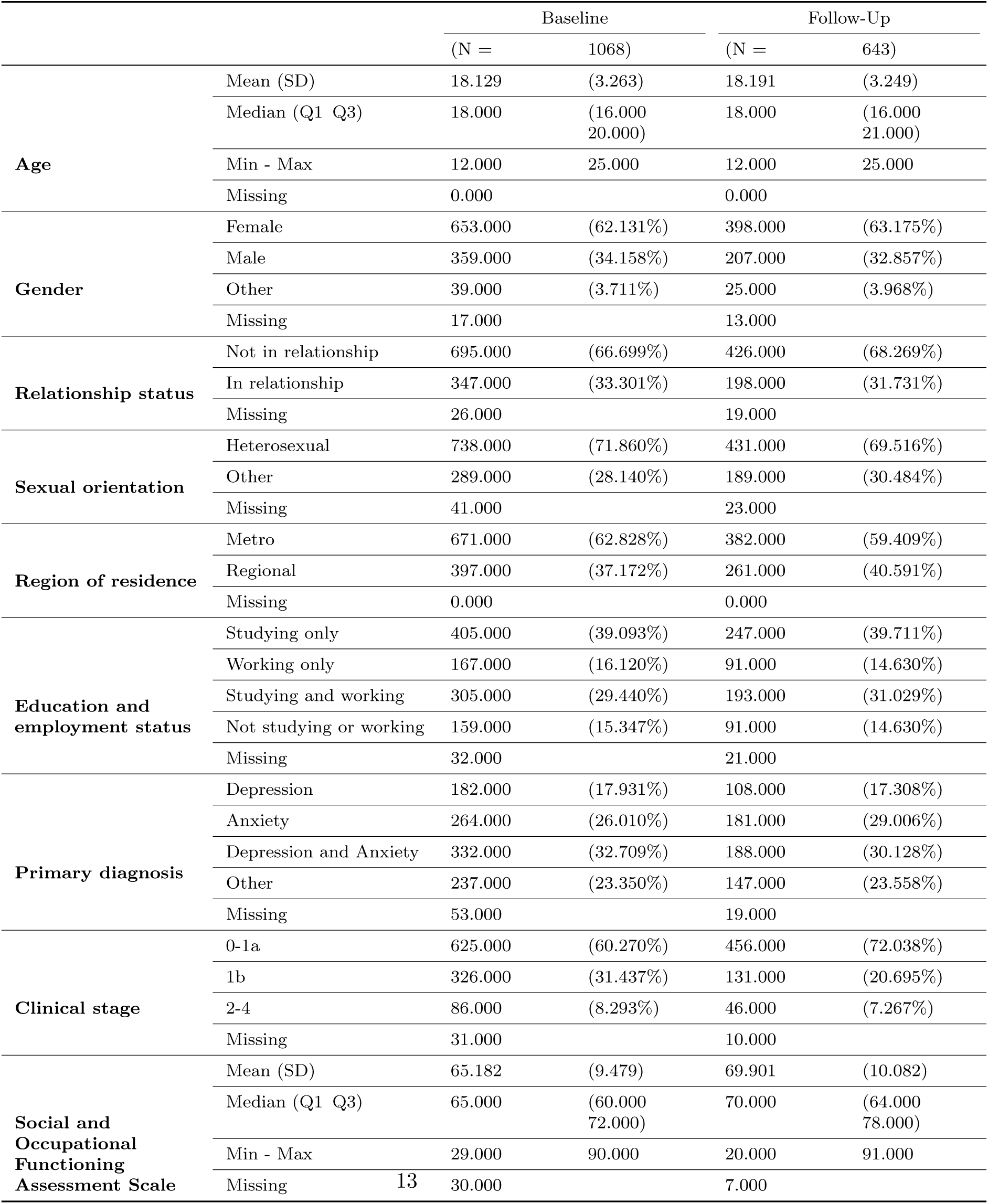
Participant characteristics.

There were 643 participants (60.205%) who completed AQoL-6D questions at the follow-up survey.

### 3.2 AQoL-6D and candidate predictors

Distributions of AQoL-6D total utility score and sub-domain scores are displayed in Figure 1. The mean utility score at baseline is 0.589 (SD = 0.235) and is 0.678 (SD = 0.238) at follow-up. Distributions of candidate predictors, K6, BADS, PHQ-9, GAD-7, OASIS and SCARED, are summarised in Table 2. PHQ-9 was found to have the highest correlation with utility score both at baseline and follow-up followed by OASIS and BADS; baseline and follow-up SCARED was found to have the lowest correlation coefficients with utility score, although all correlation coefficients can be characterised as strong.

**Fig. 1.**
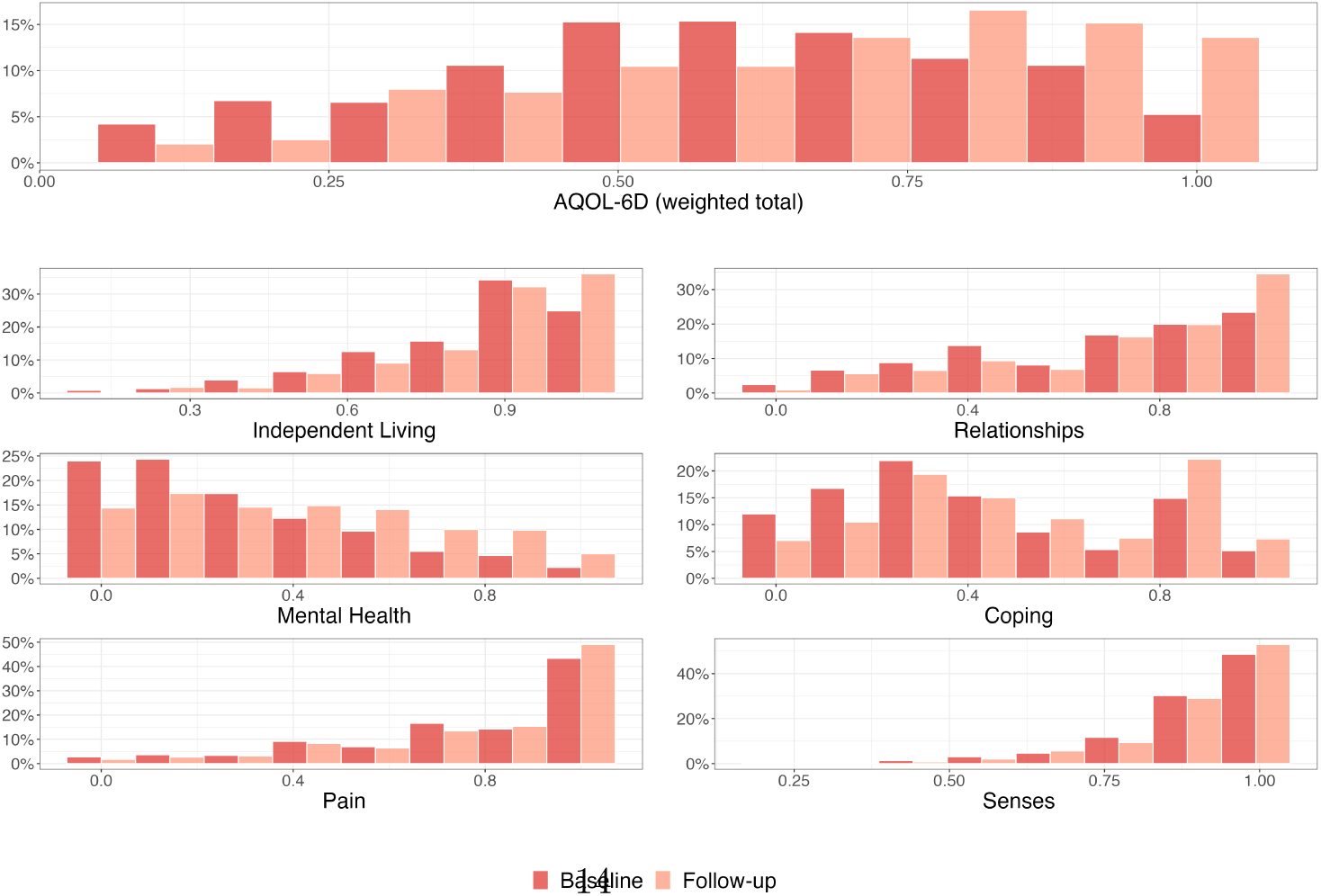
Distribution of AQoL-6D domains.

**Table 2.** Candidate predictors distribution parameters and correlations with AQoL-6D utility.

|  |  | Baseline |  | Follow-Up |  | *p* |
| --- | --- | --- | --- | --- | --- | --- |
|  |  | (N = | 1068) | (N = | 643) |  |
| **Behavioural Activation for Depression Scale (0-150)** | Mean (SD) | 78.158 | (24.823) | 89.360 | (24.435) | 0.000 |
|  | Missing | 10.000 |  | 2.000 |  | 0.000 |
|  | Correlation with AQOL-6D | 0.665 |  | 0.659 |  | 0.000, 0.000 |
| **Generalised Anxiety Disorder Scale (0-21)** | Mean (SD) | 10.382 | (5.658) | 7.945 | (5.460) | 0.000 |
|  | Missing | 6.000 |  | 2.000 |  | 0.000 |
|  | Correlation with AQOL-6D | -0.653 |  | -0.685 |  | 0.000, 0.000 |
| **Kessler Psychological Distress Scale (6 Dimension) (0-24)** | Mean (SD) | 12.156 | (5.765) | 9.811 | (5.873) | 0.000 |
|  | Missing | 4.000 |  | 2.000 |  | 0.000 |
|  | Correlation with AQOL-6D | -0.632 |  | -0.634 |  | 0.000, 0.000 |
| **Overall Anxiety Severity and Impairment Scale (0-20)** | Mean (SD) | 8.059 | (4.718) | 6.287 | (4.337) | 0.000 |
|  | Missing | 7.000 |  | 1.000 |  | 0.000 |
|  | Correlation with AQOL-6D | -0.688 |  | -0.708 |  | 0.000, 0.000 |
| **Patient Health Questionnaire (0-27)** | Mean (SD) | 12.838 | (6.616) | 9.835 | (6.484) | 0.000 |
|  | Missing | 4.000 |  | 5.000 |  | 0.000 |
|  | Correlation with AQOL-6D | -0.742 |  | -0.775 |  | 0.000, 0.000 |
| **Screen for Child Anxiety Related Disorders (0-82)** | Mean (SD) | 34.238 | (17.852) | 28.825 | (17.826) | 0.000 |
|  | Missing | 7.000 |  | 2.000 |  | 0.000 |
|  | Correlation with AQOL-6D | -0.635 |  | -0.629 |  | 0.000, 0.000 |

### 3.3 Cross-sectional models

The 10-fold cross-validated model fitting indices from mapping models using PHQ-9 are reported in Online Supplement Table A.1. There were convergence issues with beta regression models and, as they do not show major advantages in predictive performance, these models were not considered further.

Model diagnoses (such as heteroscedasticity, residual normality) suggested better model fit of the clog-log transformed OLS model, as the distribution of clog-log transformed utility are closest to normal distribution among all transformation methods. Another benefit of the clog-log transformed model is that the predicted utility score will be constrained with an upper bound of 1, thus preventing out of range prediction. Therefore, OLS with clog-log transformation was chosen as the best model for further evaluation. The GLM with Gaussian distribution and log link is commonly used in mapping studies [23], so was included to provide comparisons with other published work.

PHQ9 had the highest predictive ability followed by OASIS, BADS, GAD7 and K6 (Online Supplement Table A.2). SCARED had the least predictive capability. The confounding effect of other participant characteristics were also evaluated, with SOFAS found to independently predict utility scores in models for all six candidate predictors (*p<0.005*). Sex at birth was found to be a confounder for the K6 model (*p<0.01*). A few other confounders, including primary diagnosis, clinical staging and age were identified as weakly associated with utility in mapping models using anxiety and depression measurements other than PHQ-9. Many of these factors are unlikely to change over three months, so were not evaluated in the mixed effect models.

### 3.4 Longitudinal models

Regression coefficients of the baseline score and score changes (from baseline to follow-up) estimated in individual GLMM and LMM models are summarised in Table Table 3). In GLMM and LMM models, the prediction models using OASIS and PHQ-9 respectively had the highest R^2^ (0.681 and 0.762). R^2^ was between 0.581 and 0.681 for all GLMMs and between 0.712 and 0.762 for all LMMs. Variance of the random intercept was comparable with the residual variance.

**Table 3.** Estimated coefficients from longitudinal mapping models.

| Model | GLMM - Gaussian distribution and log link |  |  |  |  | LMM - complementary log log transformation |  |  |  |  |
| --- | --- | --- | --- | --- | --- | --- | --- | --- | --- | --- |
| Parameter | Est. | SE | CI (95%) | R2 | Sigma | Est. | SE | CI (95%) | R2 | Sigma |
| **PHQ-9** |  |  |  | **0.656** | **0.138** |  |  |  | **0.762** | **0.409** |
| SD (Intc.) | 0.114 | 0.013 | (0.09, 0.14) |  |  | 0.357 | 0.017 | (0.32, 0.39) |  |  |
| Intc. | 0.013 | 0.013 | (-0.01, 0.04) |  |  | 1.124 | 0.033 | (1.06, 1.19) |  |  |
| PHQ-9 BL | -4.497 | 0.111 | (-4.72, -4.28) |  |  | -9.623 | 0.233 | (-10.07, -9.16) |  |  |
| PHQ-9 $\Delta$ | -3.865 | 0.148 | (-4.15, -3.57) | | | -8.086 | 0.301 | (-8.69, -7.49) | | |
| **OASIS** |  |  |  | **0.681** | **0.133** |  |  |  | **0.759** | **0.411** |
| SD (Intc.) | 0.179 | 0.010 | (0.16, 0.20) |  |  | 0.432 | 0.017 | (0.40, 0.47) |  |  |
| Intc. | -0.085 | 0.015 | (-0.11, -0.06) |  |  | 0.907 | 0.033 | (0.84, 0.97) |  |  |
| OASIS BL | -5.911 | 0.183 | (-6.27, -5.55) |  |  | -12.529 | 0.363 | (-13.26, -11.82) |  |  |
| OASIS $\Delta$ | -5.613 | 0.237 | (-6.09, -5.15) | | | -12.033 | 0.475 | (-12.97, -11.09) | | |
| **BADS** |  |  |  | **0.633** | **0.143** |  |  |  | **0.732** | **0.434** |
| SD (Intc.) | 0.172 | 0.012 | (0.15, 0.20) |  |  | 0.444 | 0.018 | (0.41, 0.48) |  |  |
| Intc. | -1.388 | 0.031 | (-1.45, -1.33) |  |  | -1.891 | 0.060 | (-2.01, -1.77) |  |  |
| BADS BL | 1.064 | 0.034 | (1.00, 1.13) |  |  | 2.291 | 0.072 | (2.15, 2.43) |  |  |
| BADS $\Delta$ | 0.845 | 0.041 | (0.77, 0.93) | | | 1.850 | 0.083 | (1.69, 2.02) | | |
| **K6** |  |  |  | **0.581** | **0.153** |  |  |  | **0.712** | **0.449** |
| SD (Intc.) | 0.162 | 0.015 | (0.13, 0.19) |  |  | 0.461 | 0.018 | (0.43, 0.50) |  |  |
| Intc. | -0.033 | 0.018 | (-0.07, 0.00) |  |  | 1.034 | 0.043 | (0.95, 1.12) |  |  |
| K6 BL | -4.235 | 0.147 | (-4.52, -3.94) |  |  | -9.254 | 0.323 | (-9.90, -8.63) |  |  |
| K6 $\Delta$ | -3.554 | 0.191 | (-3.93, -3.19) | | | -7.481 | 0.368 | (-8.21, -6.78) | | |
| **SCARED** |  |  |  | **0.615** | **0.146** |  |  |  | **0.714** | **0.448** |
| SD (Intc.) | 0.181 | 0.011 | (0.16, 0.20) |  |  | 0.456 | 0.018 | (0.42, 0.49) |  |  |
| Intc. | -0.076 | 0.017 | (-0.11, -0.04) |  |  | 0.922 | 0.039 | (0.84, 1.00) |  |  |
| SCARED BL | -1.379 | 0.049 | (-1.48, -1.28) |  |  | -2.948 | 0.101 | (-3.15, -2.76) |  |  |
| SCARED $\Delta$ | -1.459 | 0.078 | (-1.61, -1.31) | | | -3.289 | 0.161 | (-3.60, -2.97) | | |
| **GAD-7** |  |  |  | **0.620** | **0.146** |  |  |  | **0.733** | **0.433** |
| SD (Intc.) | 0.160 | 0.012 | (0.13, 0.18) |  |  | 0.443 | 0.018 | (0.41, 0.48) |  |  |
| Intc. | -0.076 | 0.015 | (-0.10, -0.05) |  |  | 0.922 | 0.036 | (0.85, 0.99) |  |  |
| GAD-7 BL | -4.639 | 0.147 | (-4.93, -4.36) |  |  | -9.856 | 0.315 | (-10.48, -9.22) |  |  |
| GAD-7 $\Delta$ | -4.207 | 0.195 | (-4.58, -3.82) | | | -8.812 | 0.383 | (-9.57, -8.06) | | |

Est. - Estimate, SE - Standard Error, CI - Credible Interval, Intc. - Intercept, BL - Baseline and $\Delta$ - Change.<sup>1</sup> The BADS, GAD-7, K6, OASIS, PHQ-9 and SCARED parameters were first multiplied by 0.01.

Distribution of observed and predicted utility scores and their association from GLMM (Gaussian distribution with log link) and LMM (c-loglog transformed) using PHQ-9 are plotted in Figure 2. Compared with GLMM, the predicted utility scores from LMM converge better to the observed distribution and provide better estimations at the tail of the distribution. When the observed utility scores were low, the predicted utility were too high in the GLMM model, see Figure 2 (B). The observed and predicted distributions of utility scores for other anxiety and depression measurements were similar for LMM. However, GLM had low coverage in utility scores below 0.3 and also made predictions out of range (over 1).

**Fig. 2.**
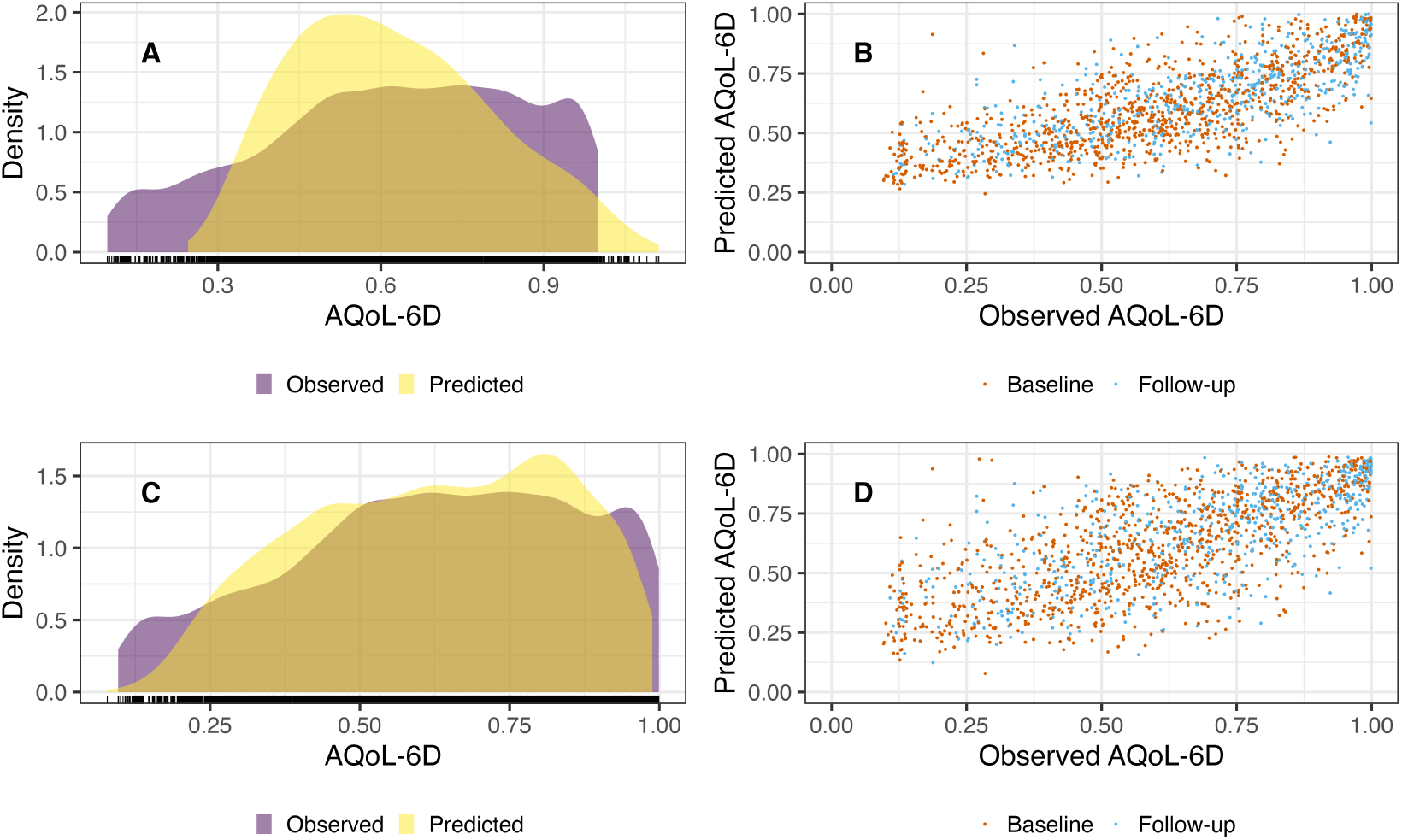
Comparison of observed and predicted AQoL-6D score from longitudinal model using PHQ-9. (A) Density plots of observed and predicted utility scores (GLMM (Gaussian distribution and log link)) (B) Scatter plots of observed and predicted utility scores by timepoint (GLMM (Gaussian distribution and log link)) (C) Density plots of observed and predicted results (LMM (complementary log log transformation)) (D) Scatter plots of observed and predicted results by timepoint (LMM (complementary log log transformation))

Model coefficients of score change from baseline were generally estimated to be lower compared with coefficients of baseline score (except for SCARED). The mean (across GLMM and LMM models) ratio of the two coefficients (*β_change_/β_baseline_*) is 0.823 for K6, between 0.802 and 0.850 for depression measurements and between 0.900 and 1.086 for anxiety measurements.

## 4 Discussion

Although there is encouraging evidence about the quality, effectiveness and cost-effectiveness of youth mental health service innovations worldwide [31, 32], the public health and economic returns from systemic reforms to support better mental health in young people needs to be better understood [33]. Our study contributes to this goal by developing tools that can facilitate derivation of QALYs from measures commonly collected in youth mental health services.

Our study is the first to evaluate longitudinal mapping ability between a wide range of affective symptom measurements and health utility in a cohort of help seeking young people. We were able to independently predict adolescent AQoL-6D from each of the six candidate measures we assessed, with PHQ-9 having the best predictive performance. Predictive performance was improved when adding a measure of functioning (SOFAS) as an additional predictor or confound to each model; SOFAS also performed well as an independent predictor. As many youth mental health services routinely collect data on at least one of the predictors included in our models, these mapping models may have widespread applicability.

Study results may be useful for helping service system planners to prioritise the measures to include in routine data collection. Although direct measurement of health utility with measures such as the ReQoL [34] may be feasible in some mental health services, relying on clinical measures that can also map to health utility may be an attractive alternative.

Our study extends recent work that showed both PHQ-9 and GAD-7 can be mapped to EQ-5D and ReQoL MAUIs [12] using data from a UK adult clinical mental health sample. Compared to that study, our mapping models have been derived from a clinical youth mental health sample, map to a different MAUI (adolescent AQoL-6D) and allow use of four additional clinical and one additional functional measures as predictors. Notably, although our sample had similar mean baseline PHQ-9 and GAD-7 scores (indicative of moderate severity) to that of the recent study, mean baseline health utility was lower (0.589 compared to 0.653 – 0.778), suggesting that the choice of instrument to map to has the potential to significantly influence the results of economic evaluations. Unlike the prior study, we did not use mixture models, but as health utility in our sample was more normally distributed, this is unlikely to be a limitation. Our modelling diagnostics suggest high performance.

A key feature of QALYs is their longitudinal dimension – health utilities are weighted and aggregated based on the time spent in varying health states. Our results suggest that cross-sectional variations in psychological distress, depression and anxiety measurements can be used to approximate the longitudinal change in health utility in this cohort. However, for psychological distress and depression measures at least, mapping algorithms developed from cross-sectional data may slightly over-estimate these changes, introducing bias into QALY predictions (overestimating QALYs for populations whose health utility improves over time, underestimating QALYs for those with deteriorating mental health).

Key strengths of our study include the novelty of our clinical youth mental health study sample, the use of clinically relevant and frequently collected outcome measures as predictors, the appropriateness and range of statistical methods deployed, the comparison of within-person and between-person differences in health utility weight predictions and highly replicable, publicly disseminated study algorithms. We acknowledge limitations that our data pertained to a single country, and we explored only one MAUI-derived utility weight. However, using utility weight input data derived from the same country as that to which an analysis pertains may be relatively unimportant [35], particularly when the MAUI is well suited to the relevant health condition (as is the case with AQoL and mental health [6]). We did not examine some potential predictors that may be more common in some mental health services (for example we explored K6, as opposed to the expanded, and commonly used measure, the K10). We also did not develop age-group specific models which would be a potentially useful extension of this work.

## 5 Conclusions

We have found that it is possible to predict both within-person and between-person differences in adolescent AQOL-6D utility weights from measures routinely collected in youth mental health services. Mapping algorithms developed from cross-sectional data can approximate longitudinal changes in health utility, but may slightly over-estimate these changes. The mapping algorithms we have developed can enable greater use of CUAs in economic evaluations using youth mental health data collections.

## Availability of data and materials

Utility mapping models, instructions on how to apply them and study replication code are distributed as part of the study data repository (https://dataverse.harvard. edu/privateurl.xhtml?token=a437cc9c-b809-4513-bbef-a2333c1c934a). R packages are available for replicating and transferring study methods (https://ready4-dev.github. io/TTU/) and applying the mapping models to out of sample data (https:// ready4-dev.github.io/youthu/).

## Ethics

The study was reviewed and granted approval by the University of Melbourne’s Human Research Ethics Committee and the local Human Ethics and Advisory Group (1645367.1)

## Data Availability

Detailed results in the form of catalogues of the utility mapping models produced by this study and other supporting information are available in the results repository https://doi.org/10.7910/DVN/DKDIB0. Tools for finding and using the utility mapping models with new prediction datasets are available as part of the youthu R package (https://ready4-dev.github.io/youthu/). The TTU (https://ready4-dev.github.io/TTU/) has tools for both replicating the study and generalising our algorithms to develop utility mapping algorithms with other utility measures and predictors. A program to replicate all steps in the study from data ingest to manuscript creation is available at https://doi.org/10.5281/zenodo.6116077.

## References

[1] MacKillop E, Sheard S. Quantifying life: Understanding the history of Quality-Adjusted Life-Years (QALYs) [Journal Article]. Social Science & Medicine. 2018;211:359–366. 10.1016/j.socscimed.2018.07.004.

[2] Neumann PJ, Goldie SJ, Weinstein MC. Preference-Based Measures in Economic Evaluation in Health Care [Journal Article]. Annual Review of Public Health. 2000;21(1):587–611. 10.1146/annurev.publhealth.21.1.587.

[3] Mortimer D, Segal L. Comparing the incomparable? A systematic review of competing techniques for converting descriptive measures of health status into QALY-weights [Journal Article]. Medical decision making. 2008;28(1):66–89.

[4] Wailoo AJ, Hernandez-Alava M, Manca A, Mejia A, Ray J, Crawford B, et al. Mapping to Estimate Health-State Utility from Non-Preference-Based Outcome Measures: An ISPOR Good Practices for Outcomes Research Task Force Report. Value in health: the journal of the International Society for Pharmacoeconomics and Outcomes Research. 2017 Jan;20(1):18–27. 10.1016/j.jval.2016.11.006.

[5] Richardson J, Iezzi A, Khan MA, Maxwell A. Validity and Reliability of the Assessment of Quality of Life (AQoL)-8D Multi-Attribute Utility Instrument. The Patient – Patient-Centered Outcomes Research. 2014 Mar;7(1):85–96. 10.1007/s40271-013-0036-x.

[6] Engel L, Chen G, Richardson J, Mihalopoulos C. The impact of depression on health-related quality of life and wellbeing: identifying important dimensions and assessing their inclusion in multi-attribute utility instruments [Journal Article]. Qual Life Res. 2018;27(11):2873–2884. 10.1007/s11136-018-1936-y.

[7] Mihalopoulos C, Chen G, Iezzi A, Khan MA, Richardson J. Assessing outcomes for cost-utility analysis in depression: comparison of five multi-attribute utility instruments with two depression-specific outcome measures [Journal Article]. The British Journal of Psychiatry. 2014;205(5):390–397.

[8] Kessler RC, Andrews G, Colpe LJ, Hiripi E, Mroczek DK, Normand SLT, et al. Short screening scales to monitor population prevalences and trends in non-specific psychological distress [Journal Article]. Psychological Medicine. 2002;32(6):959–976. 10.1017/s0033291702006074.

[9] Henry JD, Crawford JR. The short-form version of the Depression Anxiety Stress Scales (DASS-21): Construct validity and normative data in a large non-clinical sample. British journal of clinical psychology. 2005;44(2):227–239. 10.1348/014466505X29657.

[10] Spitzer RL, Kroenke K, Williams JB, Lowe B. A brief measure for assessing generalised anxiety disorder: the GAD-7. [Journal Article]. Archives of Internal Medicine. 2006;166:1092–1097.

[11] Kroenke K, Spitzer RL, Williams JB. The PHQ: validity of a brief depression severity measure [Journal Article]. Journal of general internal medicine. 2001;16(9):606–613.

[12] Franklin M, Hernández Alava M. Enabling QALY estimation in mental health trials and care settings: mapping from the PHQ-9 and GAD-7 to the ReQoL-UI or EQ-5D-5L using mixture models. Quality of Life Research. 2023;p. 1–16.

[13] Furber G, Segal L, Leach M, Cocks J. Mapping scores from the Strengths and Difficulties Questionnaire (SDQ) to preference-based utility values. Qual Life Res. 2014 Mar;23(2):403–411.

[14] Hamilton MP, Gao CX, Wiesner G, Filia KM, Menssink JM, Plencnerova P, et al.: A prototype software framework for transparent, reusable and updatable computational health economic models. Available from: https://arxiv.org/abs/2310.14138.

[15] Filia K, Rickwood D, Menssink J, Gao CX, Hetrick S, Parker A, et al. Clinical and functional characteristics of a subsample of young people presenting for primary mental healthcare at headspace services across Australia [Journal Article]. Soc Psychiatry Psychiatr Epidemiol. 2021;1433–9285 Filia, K Orcid: 0000-0001-6581-5890 Rickwood, D Menssink, J Gao, C X Hetrick, S Parker, A Hamilton, M Hickie, I Herrman, H Telford, N Sharmin, S McGorry, P Cotton, S APP1076940/National Health and Medical Research Council, Australia/ Journal Article Germany Soc Psychiatry Psychiatr Epidemiol. 2021 Jan 16. doi: 10.1007/s00127-020-02020-6.. 10.1007/s00127-020-02020-6.

[16] Richardson JR, Peacock SJ, Hawthorne G, Iezzi A, Elsworth G, Day NA. Construction of the descriptive system for the assessment of quality of life AQoL-6D utility instrument [Journal Article]. Health and quality of life outcomes. 2012;10(1):38.

[17] Cotton SM, Hamilton MP, Filia K, Menssink JM, Engel L, Mihalopoulos C, et al. Heterogeneity of quality of life in young people attending primary mental health services. Epidemiology and Psychiatric Sciences. 2022;31:e55. 10.1017/S2045796022000427.

[18] Kanter JW, Mulick PS, Busch AM, Berlin KS, Martell CR. The Behavioral Activation for Depression Scale (BADS): Psychometric Properties and Factor Structure [Journal Article]. Journal of Psychopathology and Behavioral Assessment. 2006;29(3):191–202. 10.1007/s10862-006-9038-5.

[19] Birmaher B, Brent DA, Chiappetta L, Bridge J, Monga S, Baugher M. Psychometric properties of the Screen for Child Anxiety Related Emotional Disorders (SCARED): a replication study [Journal Article]. Journal of the American Academy of Child & Adolescent Psychiatry. 1999;38(10):1230–1236.

[20] Norman SB, Cissell SH, Means-Christensen AJ, Stein MB. Development and validation of an Overall Anxiety Severity And Impairment Scale (OASIS) [Journal Article]. Depress Anxiety. 2006;23(4):245–9. 10.1002/da.20182.

[21] McGorry PD, Hickie IB, Yung AR, Pantelis C, Jackson HJ. Clinical staging of psychiatric disorders: a heuristic framework for choosing earlier, safer and more effective interventions [Journal Article]. Aust N Z J Psychiatry. 2006;40(8):616–22. 10.1111/j.1440-1614.2006.01860.x.

[22] Goldman HH, Skodol AE, Lave TR. Revising axis V for DSM-IV: a review of measures of social functioning [Journal Article]. Am J Psychiatry. 1992;149:9.

[23] Mukuria C, Rowen D, Harnan S, Rawdin A, Wong R, Ara R, et al. An Updated Systematic Review of Studies Mapping (or Cross-Walking) Measures of Health-Related Quality of Life to Generic Preference-Based Measures to Generate Utility Values. Applied Health Economics and Health Policy. 2019 Jun;17(3):295–313. 10.1007/s40258-019-00467-6.

[24] Dobson AJ, Barnett AG. An introduction to generalized linear models. CRC press; 2018.

[25] Hunger M, Baumert J, Holle R. Analysis of SF-6D index data: is beta regression appropriate? Value in health: the journal of the International Society for Pharmacoeconomics and Outcomes Research. 2011 Jul;14(5):759–767. 10.1016/j.jval.2010.12.009.

[26] Hastie T, Tibshirani R, Friedman J. The elements of statistical learning: data mining, inference, and prediction. Springer Science & Business Media; 2009.

[27] Kohavi R. A study of cross-validation and bootstrap for accuracy estimation and model selection. In: Ijcai. vol. 14. Montreal, Canada;. p. 1137–1145.

[28] Bolker BM, Brooks ME, Clark CJ, Geange SW, Poulsen JR, Stevens MHH, et al. Generalized linear mixed models: a practical guide for ecology and evolution. Trends in ecology & evolution. 2009;24(3):127–135.

[29] Gelman A, Goodrich B, Gabry J, Vehtari A. R-squared for Bayesian Regression Models [Journal Article]. The American Statistician. 2019;73(3):307–309. 10.1080/00031305.2018.1549100.

[30] R Core Team.: R: A Language and Environment for Statistical Computing. Vienna, Austria. Available from: https://www.R-project.org/.

[31] Hetrick SE, Bailey AP, Smith KE, Malla A, Mathias S, Singh SP, et al. Integrated (one-stop shop) youth health care: best available evidence and future directions [Journal Article]. Med J Aust. 2017;207(10):S5–S18. 10.5694/mja17.00694.

[32] Hamilton MP, Hetrick SE, Mihalopoulos C, Baker D, Browne V, Chanen AM, et al. Identifying attributes of care that may improve cost-effectiveness in the youth mental health service system [Journal Article]. Med J Aust. 2017;207(10):S27–S37. 10.5694/mja17.00972.

[33] Alegŕıa M, NeMoyer A, Falgàs Bagúe I, Wang Y, Alvarez K. Social Determinants of Mental Health: Where We Are and Where We Need to Go [Journal Article]. Current Psychiatry Reports. 2018;20(11):95–95. 10.1007/s11920-018-0969-9.

[34] Keetharuth AD, Rowen D, Bjorner JB, Brazier J. Estimating a Preference-Based Index for Mental Health From the Recovering Quality of Life Measure: Valuation of Recovering Quality of Life Utility Index. Value in Health. 2021;24(2):281–290. 10.1016/j.jval.2020.10.012.

[35] Khan M, Richardson J. Is the Validity of Cost Utility Analysis Improved When Utility is Measured by an Instrument with ‘Home-Country’ Weights? Evidence from Six Western Countries. Social Indicators Research. 2019 08;145. 10.1007/s11205-019-02094-z.

