## Supplementary material for "Mapping psychological distress, depression and anxiety measures to adolescent AQoL-6D utility using data from a sample of young people presenting to primary mental health services": Online supplement

### A Additional tables

Table A.1: 10-fold cross-validated model fitting index for different GLM and OLS models using PHQ-9 as predictor with the baseline data

| | Training model fit | | | Testing model fit | | |
|---|---|---|---|---|---|---|
| Model | R2 | RMSE | MAE | R2 | RMSE | MAE |
| **OLS** | | | | | | |
| **No transformation** | 0.551 | 0.157 | 0.124 | 0.545 | 0.157 | 0.124 |
| **Complementary log log transformation** | 0.542 | 0.159 | 0.123 | 0.536 | 0.159 | 0.124 |
| **Logit transformation** | 0.519 | 0.163 | 0.124 | 0.513 | 0.162 | 0.124 |
| **Log transformation** | 0.501 | 0.166 | 0.130 | 0.493 | 0.166 | 0.131 |
| **Log log transformation** | 0.484 | 0.169 | 0.128 | 0.477 | 0.168 | 0.128 |
| **GLM** | | | | | | |
| **Gaussian distribution and log link** | 0.536 | 0.160 | 0.126 | 0.530 | 0.160 | 0.126 |
| **Beta distribution and complementary log log link** | 0.551 | 0.157 | 0.124 | 0.545 | 0.157 | 0.124 |
| **Beta distribution and logit link** | 0.548 | 0.158 | 0.124 | 0.543 | 0.158 | 0.124 |

Results are averaged over ten folds. RMSE: Root Mean Squared Error; MAE: Mean Absolute Error

Table A.2: 10-fold cross-validated model fitting index for different candidate predictors estimated using GLM with Gaussian distribution and log link with the baseline data

| | Training model fit | | | Testing model fit | | |
|---|---|---|---|---|---|---|
| Model | R2 | RMSE | MAE | R2 | RMSE | MAE |
| **PHQ-9** | 0.536 | 0.160 | 0.126 | 0.530 | 0.160 | 0.126 |
| **OASIS** | 0.450 | 0.174 | 0.139 | 0.450 | 0.174 | 0.139 |
| **BADS** | 0.434 | 0.177 | 0.143 | 0.431 | 0.177 | 0.144 |
| **GAD-7** | 0.428 | 0.178 | 0.144 | 0.425 | 0.177 | 0.144 |
| **K6** | 0.397 | 0.182 | 0.143 | 0.395 | 0.182 | 0.143 |
| **SCARED** | 0.397 | 0.182 | 0.147 | 0.394 | 0.182 | 0.147 |

Results are averaged over ten folds. RMSE: Root Mean Squared Error; MAE: Mean Absolute Error

Table A.3: Estimated coefficients from utility mapping models based on individual candidate predictors with Social and Occupational Functioning Assessment Scale using GLMM (Gaussian distribution and log link)

| Parameter | Estimate | SE | CI (95%) | R2 | Sigma |
|---|---|---|---|---|---|
| **PHQ-9 SOFAS model** | | | | **0.656** | **0.139** |
| SD (Intercept) | 0.107 | 0.014 | 0.08, 0.13 | | |
| Intercept | -0.291 | 0.058 | -0.41, -0.17 | | |
| PHQ-9 baseline | -4.268 | 0.115 | -4.50, -4.04 | | |
| PHQ-9 change | -3.570 | 0.167 | -3.90, -3.24 | | |
| SOFAS baseline | 0.421 | 0.078 | 0.27, 0.57 | | |
| SOFAS change | 0.385 | 0.112 | 0.16, 0.61 | | |
| **OASIS SOFAS model** | | | | **0.684** | **0.133** |
| SD (Intercept) | 0.158 | 0.011 | 0.14, 0.18 | | |
| Intercept | -0.665 | 0.062 | -0.78, -0.54 | | |
| OASIS baseline | -5.468 | 0.179 | -5.82, -5.12 | | |
| OASIS change | -5.076 | 0.256 | -5.59, -4.57 | | |
| SOFAS baseline | 0.826 | 0.086 | 0.65, 0.99 | | |
| SOFAS change | 0.655 | 0.106 | 0.45, 0.86 | | |
| **BADS SOFAS model** | | | | **0.642** | **0.141** |
| SD (Intercept) | 0.171 | 0.011 | 0.15, 0.19 | | |
| Intercept | -1.681 | 0.060 | -1.80, -1.56 | | |
| BADS baseline | 0.966 | 0.037 | 0.89, 1.04 | | |
| BADS change | 0.736 | 0.046 | 0.64, 0.83 | | |
| SOFAS baseline | 0.561 | 0.095 | 0.38, 0.75 | | |
| SOFAS change | 0.597 | 0.120 | 0.36, 0.84 | | |
| **K6 SOFAS model** | | | | **0.590** | **0.152** |
| SD (Intercept) | 0.149 | 0.016 | 0.12, 0.18 | | |
| Intercept | -0.645 | 0.068 | -0.78, -0.51 | | |
| K6 baseline | -3.742 | 0.150 | -4.04, -3.45 | | |
| K6 change | -3.036 | 0.200 | -3.43, -2.65 | | |
| SOFAS baseline | 0.834 | 0.091 | 0.66, 1.01 | | |
| SOFAS change | 0.765 | 0.122 | 0.53, 1.00 | | |
| **SCARED SOFAS model** | | | | **0.630** | **0.144** |
| SD (Intercept) | 0.162 | 0.012 | 0.14, 0.18 | | |
| Intercept | -0.782 | 0.065 | -0.91, -0.65 | | |
| SCARED baseline | -1.238 | 0.048 | -1.33, -1.14 | | |
| SCARED change | -1.214 | 0.081 | -1.37, -1.06 | | |
| SOFAS baseline | 0.995 | 0.089 | 0.82, 1.17 | | |

Table A.3: Estimated coefficients from utility mapping models based on individual candidate predictors with Social and Occupational Functioning Assessment Scale using GLMM (Gaussian distribution and log link) *(continued)*

| Parameter | Estimate | SE | CI (95%) | R2 | Sigma |
|---|---|---|---|---|---|
| SOFAS change | 0.961 | 0.115 | 0.74, 1.19 | | |
| **GAD-7 SOFAS model** | | | | **0.625** | **0.145** |
| SD (Intercept) | 0.137 | 0.013 | 0.11, 0.16 | | |
| Intercept | -0.753 | 0.061 | -0.88, -0.63 | | |
| GAD-7 baseline | -4.209 | 0.143 | -4.49, -3.92 | | |
| GAD-7 change | -3.669 | 0.206 | -4.07, -3.27 | | |
| SOFAS baseline | 0.960 | 0.084 | 0.80, 1.13 | | |
| SOFAS change | 0.739 | 0.116 | 0.51, 0.96 | | |

Note: The BADS, GAD-7, K6, OASIS, PHQ-9, SCARED and SOFAS parameters were first multiplied by 0.01.

Table A.4: Estimated coefficients from utility mapping models based on individual candidate predictors with Social and Occupational Functioning Assessment Scale using LMM (complementary log log transformation)

| Parameter | Estimate | SE | CI (95%) | R2 | Sigma |
|---|---|---|---|---|---|
| **PHQ-9 SOFAS model** | | | | **0.767** | **0.406** |
| SD (Intercept) | 0.348 | 0.017 | 0.31, 0.38 | | |
| Intercept | 0.428 | 0.129 | 0.17, 0.69 | | |
| PHQ-9 baseline | -9.115 | 0.249 | -9.60, -8.62 | | |
| PHQ-9 change | -7.331 | 0.339 | -8.01, -6.66 | | |
| SOFAS baseline | 0.960 | 0.172 | 0.62, 1.29 | | |
| SOFAS change | 1.146 | 0.235 | 0.67, 1.61 | | |
| **OASIS SOFAS model** | | | | **0.773** | **0.401** |
| SD (Intercept) | 0.405 | 0.017 | 0.37, 0.44 | | |
| Intercept | -0.242 | 0.134 | -0.50, 0.02 | | |
| OASIS baseline | -11.522 | 0.379 | -12.28, -10.77 | | |
| OASIS change | -10.757 | 0.499 | -11.75, -9.78 | | |
| SOFAS baseline | 1.621 | 0.183 | 1.26, 1.98 | | |
| SOFAS change | 1.693 | 0.229 | 1.25, 2.15 | | |
| **BADS SOFAS model** | | | | **0.744** | **0.427** |
| SD (Intercept) | 0.439 | 0.018 | 0.40, 0.47 | | |
| Intercept | -2.549 | 0.124 | -2.79, -2.31 | | |
| BADS baseline | 2.069 | 0.080 | 1.91, 2.22 | | |
| BADS change | 1.606 | 0.092 | 1.43, 1.79 | | |
| SOFAS baseline | 1.258 | 0.207 | 0.85, 1.66 | | |
| SOFAS change | 1.528 | 0.243 | 1.05, 2.00 | | |
| **K6 SOFAS model** | | | | **0.731** | **0.436** |
| SD (Intercept) | 0.443 | 0.018 | 0.41, 0.48 | | |
| Intercept | -0.296 | 0.152 | -0.60, 0.00 | | |
| K6 baseline | -8.161 | 0.333 | -8.80, -7.52 | | |
| K6 change | -6.364 | 0.381 | -7.12, -5.62 | | |
| SOFAS baseline | 1.805 | 0.202 | 1.41, 2.21 | | |
| SOFAS change | 1.990 | 0.244 | 1.51, 2.47 | | |
| **SCARED SOFAS model** | | | | **0.736** | **0.432** |
| SD (Intercept) | 0.419 | 0.018 | 0.38, 0.46 | | |
| Intercept | -0.615 | 0.137 | -0.88, -0.35 | | |
| SCARED baseline | -2.654 | 0.101 | -2.85, -2.45 | | |
| SCARED change | -2.774 | 0.162 | -3.09, -2.46 | | |
| SOFAS baseline | 2.169 | 0.188 | 1.80, 2.53 | | |

Table A.4: Estimated coefficients from utility mapping models based on individual candidate predictors with Social and Occupational Functioning Assessment Scale using LMM (complementary log log transformation) *(continued)*

| Parameter | Estimate | SE | CI (95%) | R2 | Sigma |
|---|---|---|---|---|---|
| SOFAS change | 2.337 | 0.236 | 1.87, 2.80 | | |
| **GAD-7 SOFAS model** | | | | **0.745** | **0.425** |
| SD (Intercept) | 0.406 | 0.018 | 0.37, 0.44 | | |
| Intercept | -0.571 | 0.132 | -0.83, -0.31 | | |
| GAD-7 baseline | -8.900 | 0.306 | -9.49, -8.29 | | |
| GAD-7 change | -7.601 | 0.398 | -8.37, -6.81 | | |
| SOFAS baseline | 2.111 | 0.182 | 1.75, 2.47 | | |
| SOFAS change | 1.866 | 0.237 | 1.41, 2.33 | | |

Note: The BADS, GAD-7, K6, OASIS, PHQ-9, SCARED and SOFAS parameters were first multiplied by 0.01.
